# Evaluation of age groupings used for syndromic surveillance

**DOI:** 10.1101/2025.01.10.25320339

**Authors:** R. A. Morbey, D. Todkill, H. E. Hughes, A. Charlett, A. J. Elliot

## Abstract

Public health surveillance stratifies populations into age groups to help identify threats and provide appropriate responses. However, there is considerable variation in the age groupings used for epidemiology both between and within countries.

We evaluate the age groups (under 1, 1-4, 5-14, 15-44, 45-64, over 65 years) used for syndromic surveillance in England. Comparing the existing age grouping with alternatives and using syndromic data to suggest new age groupings that maximise the homogeneity within groups and heterogeneity between groups. Data between November 2011 and March 2024 was extracted from four syndromic systems including 79 different syndromic indicators.

Correlations between time series for individual ages in years were used to calculate homogeneity of specific age groups and age groupings (collections of age groups that completely span 0 to 90 years). Young adolescents were identified as a specific age group with distinct trends different to younger children or older adolescents. The current age group of 5 to 14 years was found to be more heterogeneous that over age groups, even those with a much wider span. Also, the age group over 65 years was assessed to be too broad and would benefit from being split into those over 90 years and below. Thus, our recommendation is a new age grouping for syndromic surveillance consisting of under 1s, 1 to 4, 5 to 8, 9 to 17, 18 to 33, 34 to 50, 51 to 67, 68 to 89 and over 90 years.

## Introduction

Public health surveillance involves monitoring population health for emerging threats (early warning) and providing situational awareness of current health trends {Smith, 2019 #1250}. Certain diseases or health threats can often have a more serious impact on certain age groups, e.g. the elderly or infants, and therefore for epidemiologists tracking and monitoring disease trends, it is important to disaggregate surveillance data by age. This enables the identification of evidence of differential trends amongst different age groups, which is key information that enables epidemiologists and public health organisations to identify threats and provide an appropriate response (1).

Whilst it is common practice to report public health surveillance data disaggregated by age, there is no standard set of age groups, and age groupings used across different surveillance systems vary greatly. In particular, the COVID-19 pandemic highlighted how the use of different age groupings within and across countries hindered the ability to perform rapid cross-country analyses (2). Often, it is unclear why a particular set of age groups has been chosen for a surveillance system and there may be several considerations, including data availability, comparability with other systems, local reporting requirements, clinical expert opinion etc.

The UK Health Security Agency (UKHSA) is the body responsible for health protection and infectious disease surveillance in England. As part of its extensive surveillance capacity, UKHSA has six national syndromic surveillance systems that are used alongside traditional laboratory surveillance as an all-hazards public health monitoring programme. Currently the six syndromic systems use the following age groups for routine epidemiological reporting: under 1 year, 1 to 4 years, 5 to 14, 15 to 44, 45 to 64 and over 65 years. These age groups have been in place historically for many years.

The World Health Organisation (WHO) provides definitions for life stages represented by age groups for public health indicators (Table 1) (2, 3).

**Table 1.** World Health Organisation defined age groups for public health surveillance.

| Age group | Definition |
| --- | --- |
| 0-6 days | early neonates |
| 7-27 days | late neonates |
| 28-364 days | post-natal infants |
| 1-4 years | young children |
| 5-9 years | older children |
| 10-14 years | young adolescents |
| 15-19 years | older adolescents |
| 20-24 years | young adults |
| 25-59 years | adults (in five-year age groups) |
| 60-99 years | older adults (in five-year age groups) |
| 100+ | older adults |

The WHO recommendations are in general designed for using age groups with a span of five years, whilst separating infants as a distinct group. The defined names used by WHO for these age groups is a recognition of the developmental stage in a person’s life course which is one approach to defining age groups. A similar approach could use life stages such as beginning or ending full time schooling or retirement to define age groups. Whilst, these methods are useful for population surveillance individuals may vary widely in how they develop and other groups may be relevant for specific diseases (4). An alternative approach to determining age groups is to start by analysing data, for example Geifman et al (5) used a disease database to develop age groups based on clustering methods such as k-means.

In this study, we evaluate the UKHSA syndromic surveillance age grouping by comparing them to alternatives and by examining data used for syndromic surveillance across a wide range of syndromic systems and indicators.

## Methods

### Syndromic surveillance data

Anonymised surveillance data were extracted from UKHSA real-time syndromic surveillance systems: National Health Service (NHS) 111 telehealth calls (NHS 111); general practitioner (GP) out-of-hours (OOH) contacts (GPOOH); emergency department attendances (the ED Syndromic Surveillance System, EDSSS); and ambulance dispatch calls (National Ambulance Syndromic Surveillance System, NASSS). One of the UKHSA systems, an online telehealth service (‘NHS 111 online’) was excluded because it did not include children aged under 5 years. Furthermore, the GP in-hours system was excluded because age data were only available in pre-aggregated age groups that could not be further disaggregated. Thus, the data used in this study were ED attendances, ambulance dispatch calls, GP out-of-hours contacts and NHS 111 telehealth calls. For NASSS, data were only included from two regions where age stratified data were available.

Data were extracted in the form of daily counts by syndromic indicator and age in years at time of presenting to health care. Historical data were extracted up to 14 March 2024, with the earliest date available varying by system: GPOOH 5 November 2011, NASSS 1 April 2016, EDSSS 5 November 2018 and NHS 111 calls 1 October 2020. The start date for data extraction for each system was chosen to exclude pilot periods when only partial data were available. The data were cleaned to remove outliers caused by days where only partial data were available. Similarly, spikes in data caused by batch reporting and exceptional data during the peak of the COVID-19 pandemic were excluded.

A wide range of syndromic indicators are used in ongoing surveillance to provide a service that is sensitive to many potential public health threats. A few syndromic indicators designed to identify rarer events include very sparse data, particularly for some ages, e.g. ED attendances for Guillain-Barre or GP out-of-hours measles contacts. Therefore, indicators were excluded if any of the ages below 85 years only had zero counts, this left 79 indicators available for analysis (listed in appendix).

### Alternative age groupings

The existing age grouping used for syndromic surveillance by UKHSA comprises six age groups: under 1 year, 1 to 4 years, 5 to 14, 15 to 44, 45 to 64 and over 65 years. The under 1 years is the only age group that contains just a single year, because it is believed important to identify public health hazards that particularly affect infants aged under 12 months. To evaluate how distinct infants are from other ages, we compared the UKHSA syndromic age grouping with an alternate grouping with a single under 5 years age group. At the other end of the scale the literature suggests that the over 65 years age group is too broad (6), and so an alternate grouping was considered using an additional over 80 years age group.

The WHO published guidelines for health data age groups based on developmental stages which we used for comparison. The WHO age grouping we evaluated included the following age groups: infants (under 1 years), young children (1 to 4 years), older children (5 to 9), young adolescents (10 to 14), older adolescents (15 to 19), young adults (20 to 24), adults (25 to 59), and older adults (60 to 99). The WHO guidelines, suggest separating infants further into early neonates (0-6 days), late neonates (7-27 days) and post-natal infants, however we did not include any age groups smaller than a 1-year span, as this level of detail is not available in most systems.

Using a disease database, a study by Geifman et al. suggested the following alternate age grouping of nine age groups: 0 to 2 years, 3 to 5, 6 to 13, 14 to 18, 19 to 33, 34 to 48, 49 to 64, 65 to 78 and 79 to 98. These groups were chosen by using a k-means clustering approach and therefore, here this grouping is referred to as ‘k-means’ (5). Finally, for comparison as a control a ‘decile’ age grouping was included, where each age group contained ten years, e.g. 0-9 years, 10-19 etc.

### Correlation between ages

Daily counts for all 79 syndromic indicators were combined and reshaped so that each age (in years) formed a separate column of data, where the rows were the daily counts for a specific syndromic indicator from one system on a specific date. Next, correlations between different ages were calculated using all the data from every indicator. Thus, a high correlation between two ages was indicative of similar trends in counts between those two ages across all indicators.

### Comparison between age groups

To measure the similarity between ages within an age group, the mean of all pairwise correlations of ages within the age group were calculated. Thus, an age group with a higher mean correlation covers an age range which is less diverse than an age group with a lower mean correlation. The trivial example of an age group containing just one year (e.g. under 1-year olds) would have a perfect mean correlation score of 1.

It was possible to compare two alternate age groupings, both consisting of age groups that span all ages by calculating the ‘area under the groups’ (AUG). For each age group within an age grouping, the AUG is calculated as its mean pairwise correlation score multiplied by its span in years. Then the AUG for an age grouping is the sum of the AUGs of its constituent age groups.

Importantly, the AUG method of comparing age groupings makes it possible to sequentially add age breaks, creating new age groupings that minimise the differences within age groups whilst maximising the difference between age groups. Thus, age breaks were added to ungrouped data to find the best age grouping made of two age groups, then three age groups and so forth up to the best grouping with nine age groups.

Syndromic data includes data for the very elderly, but data becomes increasing sparse for patients over 90 years. Therefore correlations are much weaker for the oldest ages and ages over 90 years were excluded from the calculations of mean correlations and AUG.

## Results

We extracted daily data for 79 different syndromic indicators across the four national syndromic surveillance systems, comprising 22.9 million daily counts when disaggregated by age in years at time of presentation. Once disaggregated, over half of the daily counts were zero and the mean daily count across all indicators was 3.2 with a maximum of 1,568. For each age in years there were 210,256 observations used to compare trends.

### Correlations between ages

The pairwise correlations between each age in years were calculated and are shown as a heatmap in figure 1. As is to be expected, correlations were highest between adjacent ages, shown by the highest values being around the diagonal line in figure 1. Correlations fell markedly for ages over 95 where data is sparse, confirming our decision to exclude the very elderly from our calculations of mean correlation.

**Figure 1:**
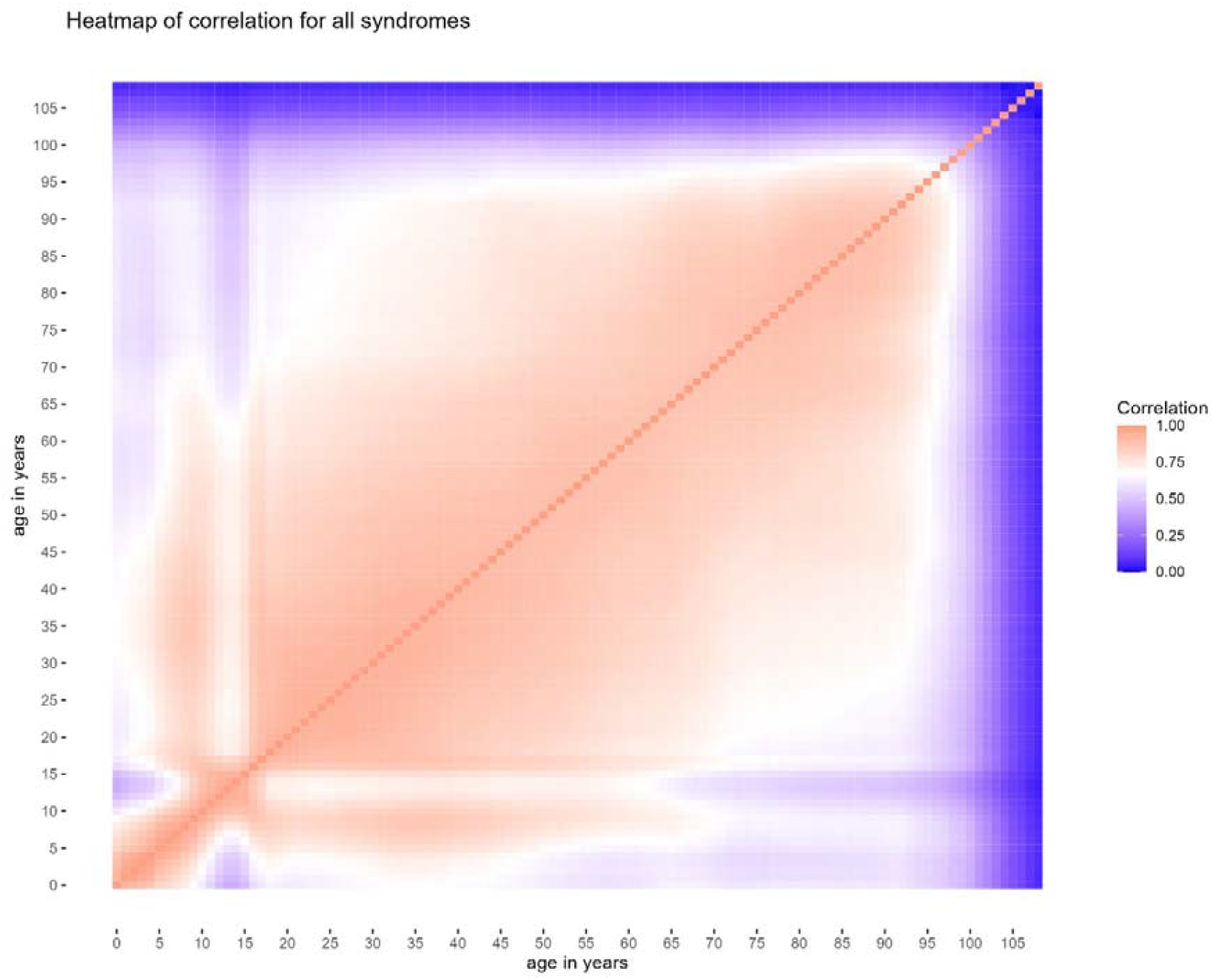
heatmap of correlation between age in years for all syndromic indicators

Interestingly, the correlation between ages was not a simple monotonic one, where each age is always more closely correlated to ages closer to itself than other ages. For example, older children were more closely correlated to young adults and adults than they were to young adolescents. Young adolescents, appeared to be a distinct group with relatively poor correlations with children and older adolescents. Infants and young children had a relatively high correlation with older children but a much lower correlation with adults. By contrast, correlations between 17 to 70 year olds appeared fairly similar as did 70 to 90 year olds.

### Comparison between age groupings

An age group that contains just one year had a perfect correlation of 1, whilst the mean pairwise correlation across all ages 0 to 90 years, i.e. the ungrouped data, was 0.771. Naturally, narrow age groups will tend to have a higher mean correlation than wider age groups, however not all age groups of the same span (i.e. number of different years in group) had the same mean correlation. For example, the two narrowest age groups used by UKHSA for syndromic surveillance had the highest mean correlations; under 1 years, and 1 to 4 years had mean correlations respectively of 1 and 0.961. However, the next narrowest of the UKHSA age groups, 5 to 14 years had the lowest mean correlation of 0.836. By contrast the adult age groups of 15 to 44, 45 to 64 and 65 to 90 had mean correlations of respectively, 0.888, 0.872, and 0.862. Table 2 shows the mean correlations of the different age groups analysed along with the area under the group calculation (mean correlation multiplied by span of age group). Similarly, within the deciles age group, where all age groups have the same span, correlations vary from 0.929 for 20 to 29 and 30 to 39 years, down to 0.860 for 10 to 19 years.

**Table 2.**
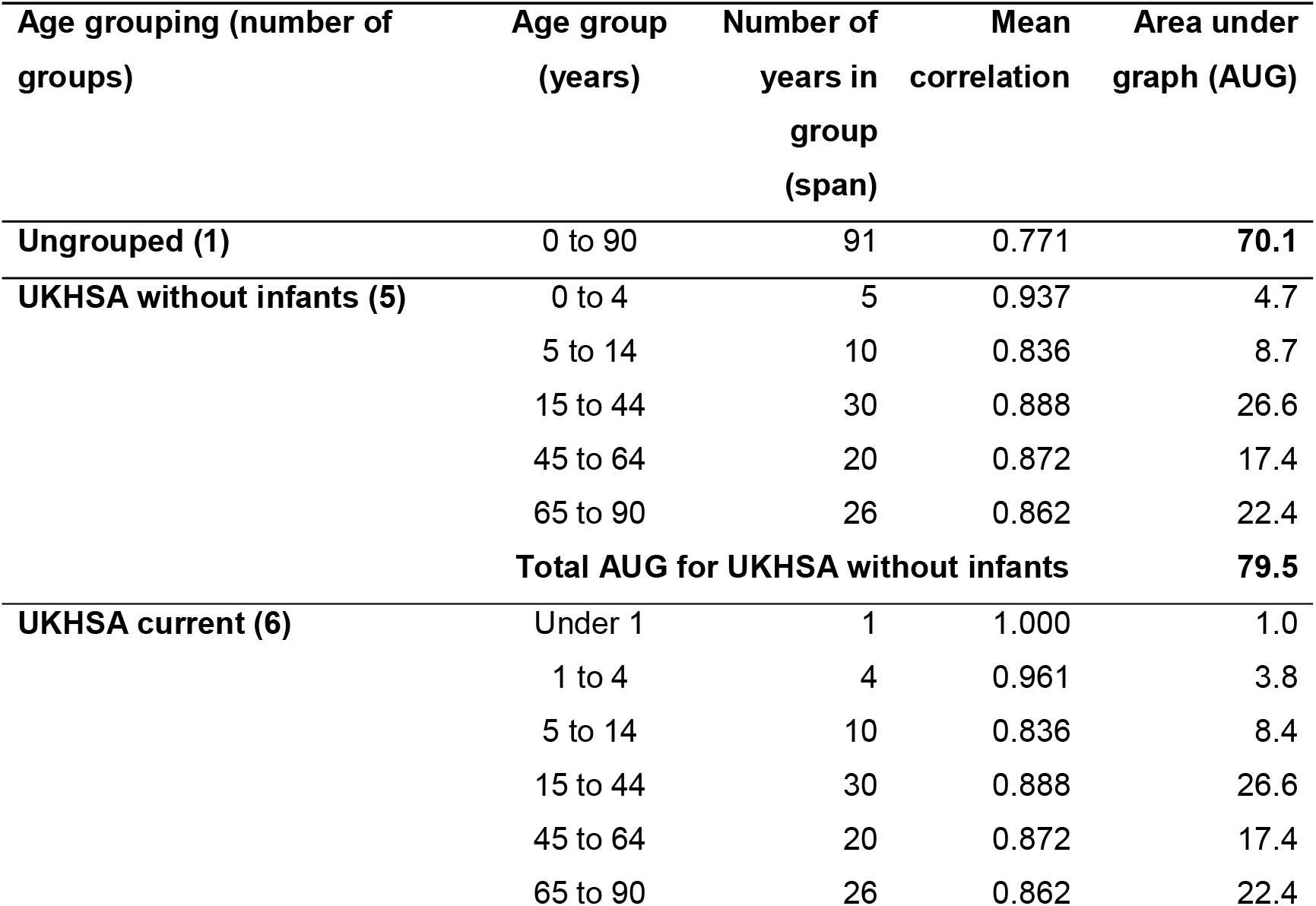
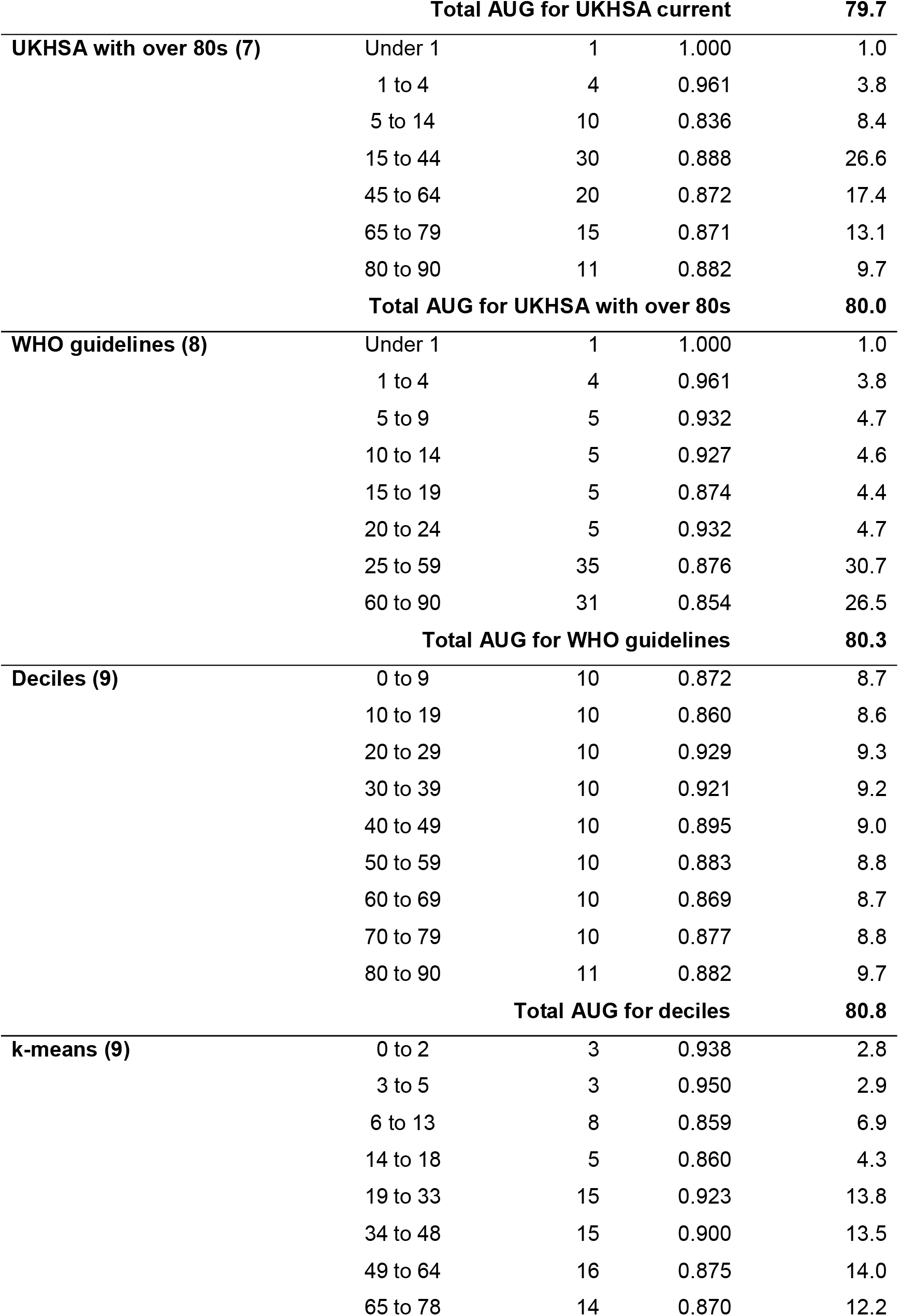
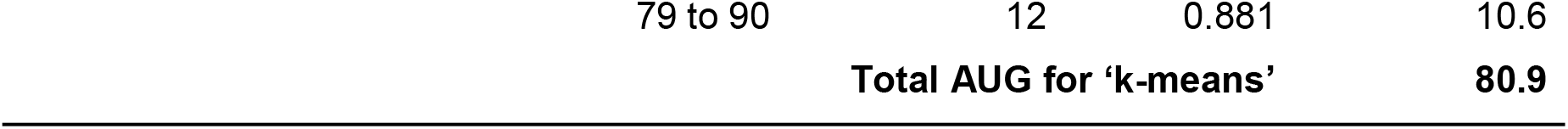
Comparison of mean correlation of ages within age groups.

All age groupings had, as expected, a higher AUG score than leaving the data ungrouped which had a score of 70.1 The current UKHSA age grouping had an AUG score of 79.8, and the grouping with the highest AUG score was k-means with 80.9. The AUG score was higher for age groupings with more age groups, and there is a theoretical maximum score of 91 if the data were split into 91 age groups each containing just 1 year. Ignoring the trivial cases of ungrouped and single year age groups, the highest mean correlations were in the 1 to 4 and 3 to 5 years, respectively 0.961 and 0.950; whilst the lowest correlations were in the 5 to 14 (0.836) and the 60 to 90 (0.854) age groups.

### New age groupings by sequentially adding age breaks

New age groupings were developed by sequentially adding age breaks to maximise the increase in the AUG score. Thus, age groupings containing 2 to 9 age groups were created. The first age break added was 50, creating two age groups of 0 to 50 and over 50 years.

Subsequently age breaks were added, in order, at 8, 17, 67, 33, 4, 80, and 10 years. Figure 2 shows how each age break increased the mean correlation.

**Figure 2:**
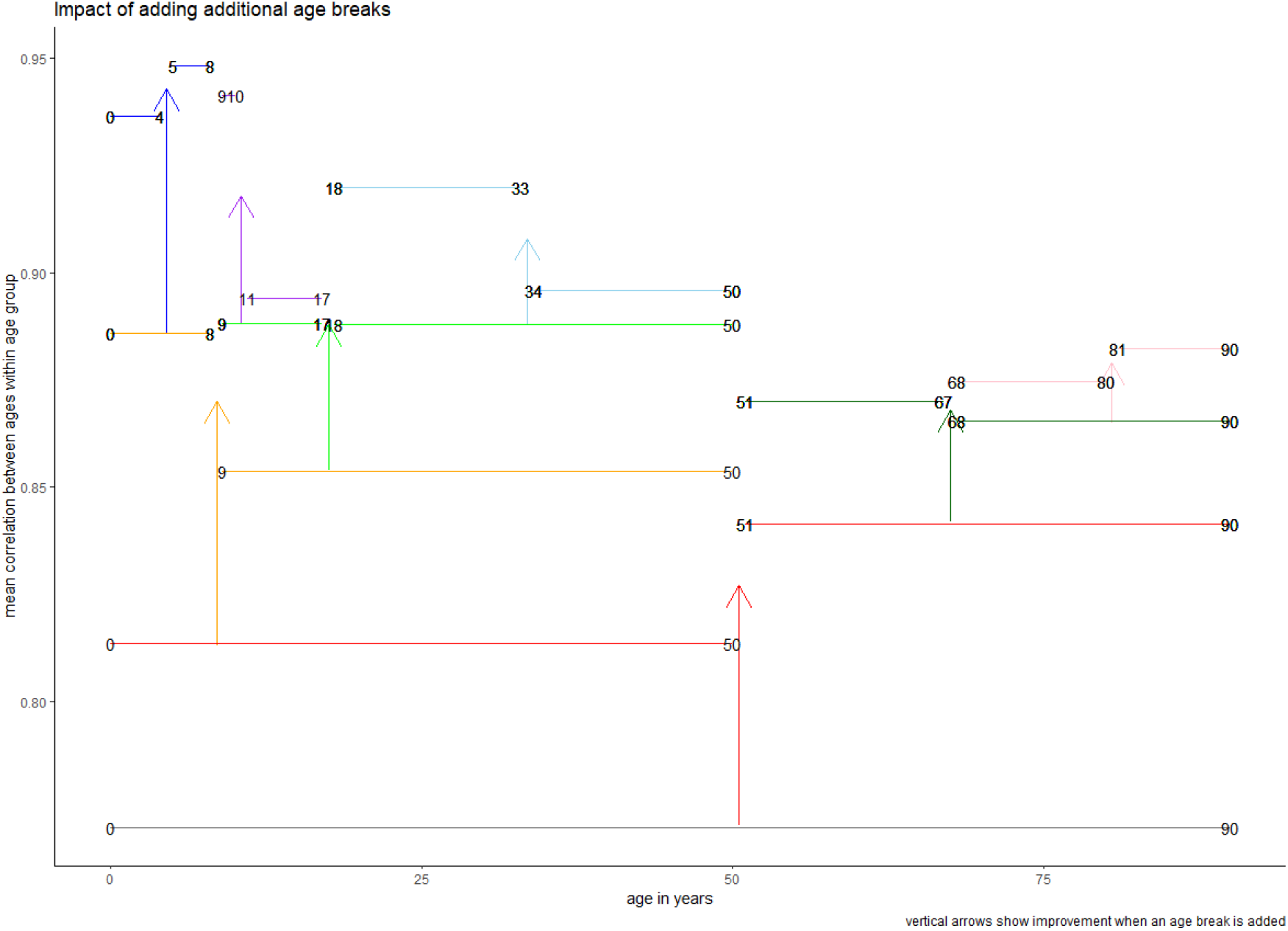
impact of adding additional age breaks (vertical arrows show improvement when an age break is added)

The first age break created two groups of similar spans, but the next two age breaks split groups into very unequal sized bands, splitting 0 to 50 years into 0 to 8 and 9 to 50 and then splitting 9 to 50 into 9 to 17 and 18 to 50. In general, narrower age bands had higher mean correlations but the 9 to 17 age group had a very similar mean correlation to the 18 to 50 age group. The next four age breaks split age groups into roughly equal spans, but the final age break created a narrow new age group of 9 to 10 year olds (Table 2).

**Table 2:**
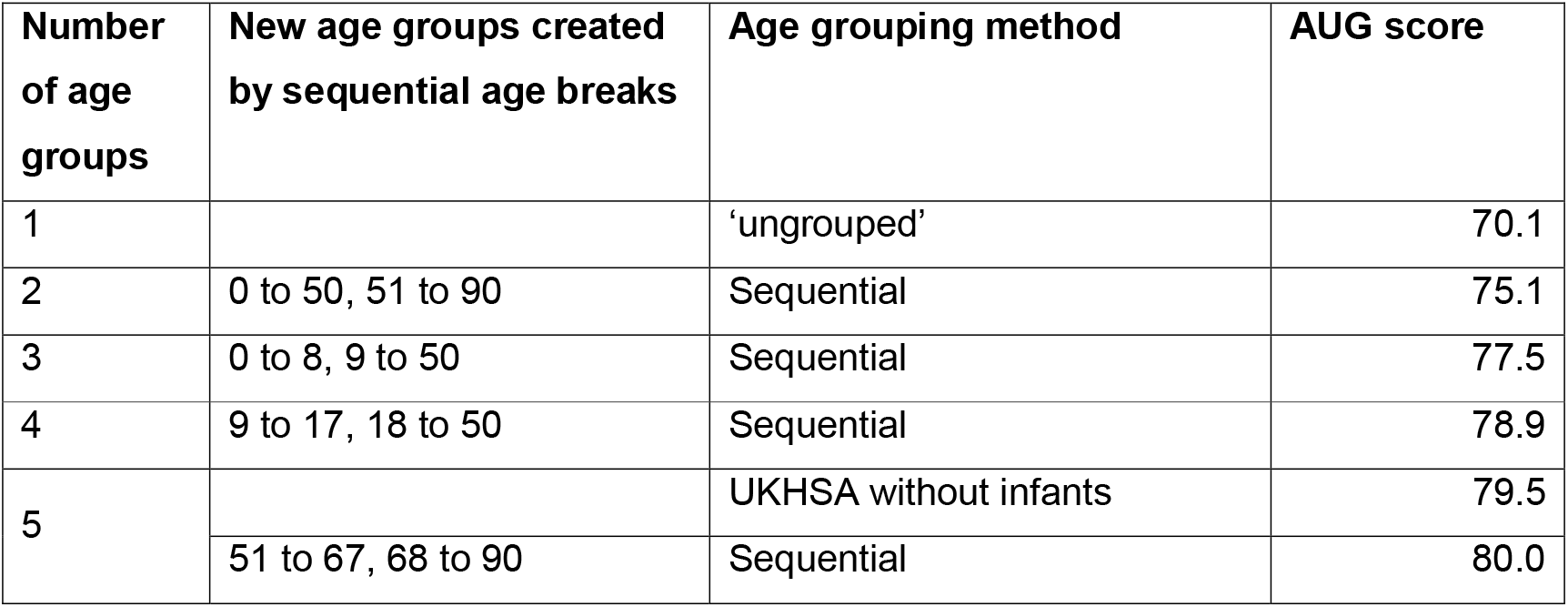
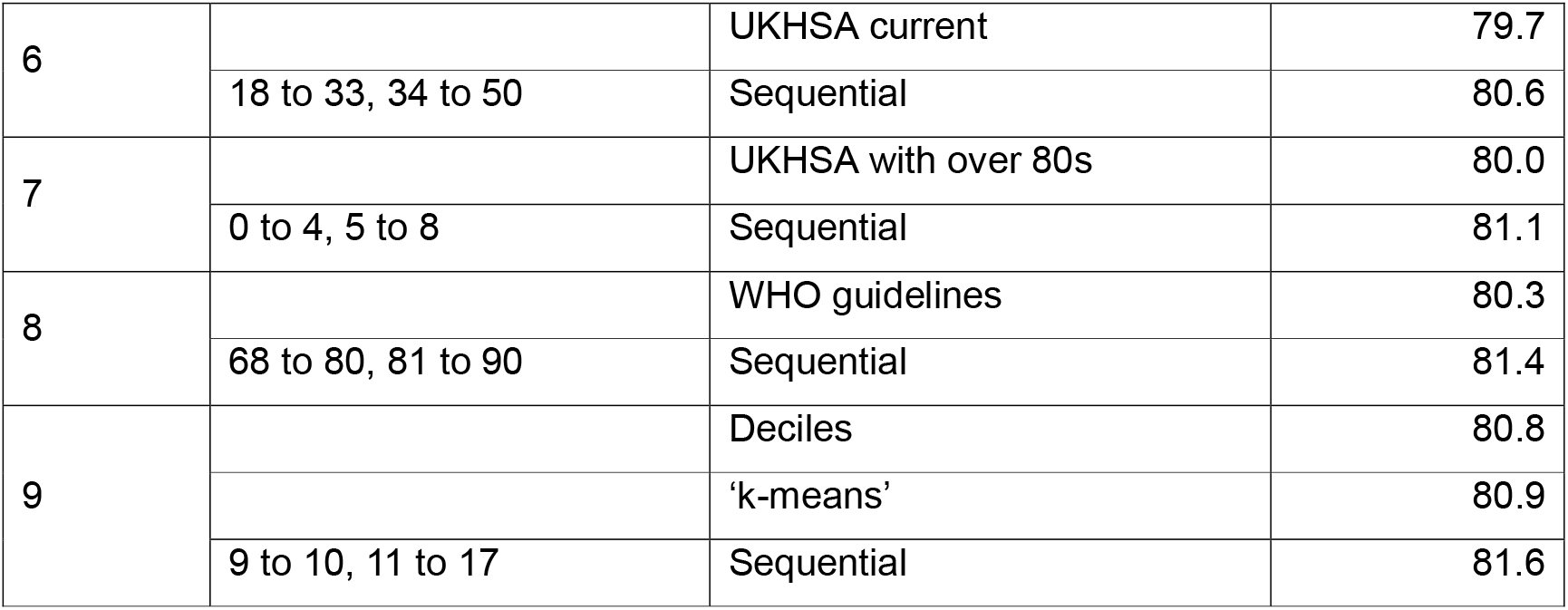
Comparison of age groupings.

The main difference between the age grouping of six groups used by UKHSA and that suggested by the sequential approach were for the younger ages. UKHSA syndromic surveillance use under 1 years, 0 to 4 years and 5 to 14 age groups whilst the sequential method for six age groups had just two child age groups, 0 to 8 and 9 to 17. When compared to the WHO guidelines based on lifestyle development, using 8 age groups the differences were that the sequential method did not suggest separating under 1 years, or splitting adolescents into younger and older but did suggest an extra age group for those aged over 80. Also, by comparison with WHO the upper bands for young adults were moved roughly 10 years later, e.g. young adults being 18 to 33 instead of 20 to 24 and adults being 34 to 50 not 25 to 59.

## Discussion

We used national syndromic surveillance data from 2011 until 2024 to compare trends in 79 different syndromic indicators across people aged from under 1 year to 90 years old. We calculated how closely correlated each age in years was to each other age and quantified how well different age groups and age groupings performed in grouping together ages with similar trends. Furthermore, we used our data to identify new age groupings that maximised the similarities within age groups and differences between age groups and compared these with existing age groupings.

Analysis of our correlation matrix suggest three distinct age groups for children, with much more homogeneity amongst adults, but with differences between those over retirement age (approximately 65 years) and younger adults. The pattern of 5 to 10 year olds being more closely correlated to adults aged under 65, than older children is reminiscent of transmission patterns for infectious disease where contacts between parents and offspring have an important role to play (7).

We confirmed findings in other literature that improvements can be made by dividing older adults into two age groups, with an age break at over 80 years representing increased frailty (6, 8). At the other end of the age scale we found that including a separate age group for infants only gave marginal improvements. However, there may be other important clinical and epidemiological reasons why infants should be considered separately, for instance their vulnerability to some infectious diseases or their immune status.

A key finding of our analysis was that the 5 to 14 years age group does not appear to be a homogonous group, with a mean correlation at 0.836, which was lower than any other group studied, even those that were much wider, e.g. 15 to 44 year olds. Amongst the deciles, the lowest mean correlation was for 10 to 19 years olds at 0.860. Splitting the 5 to 14 year age group, as in the WHO guidelines of 5 to 9 and 10 to 14 resulted in much higher mean correlations of 0.932 and 0.927 respectively.

There are many alternative ways in which age groups can be identified from data, for instance clustering methods. However, one issue with unsupervised machine learning methods like clustering is how to interpret age groups that result in overlapping or discontinuous age groups (9). Another approach is to identify the stratification that maximises the difference between groups, (10) although this is easier when considering a single primary outcome rather than a multi-hazards surveillance system.

Whilst we have concentrated on similarities between ages within syndromic data, there are other public health considerations which may make particular age groupings useful. Firstly, vaccination programmes may be tailored to specific ages (e.g. childhood immunisation schedules) and therefore it is important to distinguish these age groups in surveillance data. Secondly, public health interventions may be based on life stage development, for instance sending communications via schools, so age groups that reflect these stages will be important. Finally, some public health threats impact different age groups, so age groupings may be chosen to reflect these risks.

We have deliberately chosen to consider a very wide range of syndromic indicators, reflecting many potential public health hazards and threats, as it is practical to have a single age grouping for reporting. However, future work could consider whether there are key differences based on health care system or indicators which may suggest different age groups are appropriate for different indicators, for example respiratory illness vs impact of extreme heat. Furthermore, data could be stratified by gender or geography to identify more granular differences.

## Conclusion

A key finding from this evaluation is that for syndromic data the 5 to 14 years age group is not homogenous, therefore we would recommend not using this age group for children. The data suggests, age groups of under 5 years, 5 to 8 and 9 to 17 would be a better grouping, however there are clinical reasons why under 1 years may be retained as a distinct group. At the other end of the scale we confirmed that the over 65 years could be split into 65 to 80 and over 80s as a way of distinguishing frail and older adults. Table 3 includes our recommendations for a nine-group age grouping based on the data in our study.

**Table 3:** Recommended age grouping for syndromic surveillance (years)

| Recommendation | Existing UKHSA age grouping |
| --- | --- |
| Under 1s | Under 1s |
| 1 to 4 | 1 to 4 |
| 5 to 8 | 5 to 14 |
| 9 to 17 |  |
| 18 to 33 | 15 to 44 |
| 34 to 50 |  |
| 51 to 67 | 45 to 64 |
| 68 to 89 | Over 65 |
| Over 90 |  |

## Data Availability

All data produced in the present study are available upon reasonable request to the authors

## Appendix list of syndromic indicators included in analysis

GPOOH: acute bronchitis

GPOOH: acute respiratory infection

GPOOH: asthma

GPOOH: blood in stools

GPOOH: cardiac conditions

GPOOH: chest pain

GPOOH: chickenpox

GPOOH: diarrhoea

GPOOH: difficulty breathing, wheeze or asthma

GPOOH: double vision

GPOOH: eye problems

GPOOH: fever

GPOOH: gastroenteritis

GPOOH: gastrointestinal conditions

GPOOH: heat or sunstroke

GPOOH: hepatitis

GPOOH: hypothermia

GPOOH: impact of cold

GPOOH: impact of heat

GPOOH: influenza-like illness

GPOOH: injuries

GPOOH: insect bites

GPOOH: malaise

GPOOH: pharyngitis or scarlet fever

GPOOH: rash

GPOOH: respiratory conditions

GPOOH: stroke

GPOOH: urinary tract infection

GPOOH: vomiting

NASSS: abdominal pain

NASSS: allergic reactions

NASSS: cardiac or respiratory arrest

NASSS: chest pain

NASSS: collapsed with unknown problem

NASSS: convulsions or fitting

NASSS: difficulty breathing

NASSS: eye problems

NASSS: headache

NASSS: injuries

NASSS: other sickness

NASSS: overdose or ingestion or poisoning

NASSS: stroke

NASSS: unconscious or passing out

NASSS: carbon monoxide inhalation or poisoning

EDSSS: acute alcohol intoxication

EDSSS: acute bronchiolitis

EDSSS: acute respiratory infection

EDSSS: asthma

EDSSS: bones or joint conditions

EDSSS: burns

EDSSS: cardiac conditions

EDSSS: encephalitis

EDSSS: gastroenteritis

EDSSS: gastrointestinal conditions

EDSSS: HUS-like

EDSSS: impact of cold EDSSS: influenza-like illness

EDSSS: meningitis

EDSSS: meningococcal sepsis

EDSSS: myocardial ischaemia

EDSSS: overdose or poisoning

EDSSS: pneumonia

EDSSS: respiratory conditions

EDSSS: stroke

EDSSS: heat or sunstroke

EDSSS: covid-19-like

NHS111_calls: cold or flu

NHS111_calls: cough

NHS111_calls: diarrhoea

NHS111_calls: difficulty breathing

NHS111_calls: eye problems

NHS111_calls: fever

NHS111_calls: headache

NHS111_calls: heat exposure or sunburn

NHS111_calls: insect bites

NHS111_calls: mental health

NHS111_calls: sleep difficulties

NHS111_calls: sore throat

NHS111_calls: vomiting

## References

1. Todkill D, Lamagni T, Pebody R, Ramsay M, Woolham D, Demirjian A, et al. Persistent elevation in incidence of pneumonia in children in England, 2023/24. Eurosurveillance. 2024;29(32):2400485.

2. Diaz T, Strong KL, Cao B, Guthold R, Moran AC, Moller A-B, et al. A call for standardised age-disaggregated health data. The Lancet Healthy Longevity. 2021;2(7):e436–e43.

3. WHO. WHO Glossary of Health Data, Statistics and Public Health Indicators 2024 [Available from: https://cdn.who.int/media/docs/default-source/documents/ddi/indicatorworkinggroup/glossary_of_terms_june-2024.pdf?sfvrsn=ac699779_1.

4. Chen J, Yao L, Alamoudi AJ, Aleya L, Gu W. A Historical Misconception in Clinical Trials of Drugs for Cancer-Age Grouping. J Pers Med. 2022;12(12).

5. Geifman N, Cohen R, Rubin E. Redefining meaningful age groups in the context of disease. Age (Dordr). 2013;35(6):2357–66.

6. Vrettos I, Anagnostopoulos F, Voukelatou P, Kyvetos A, Theotoka D, Niakas D. Does Old Age Comprise Distinct Subphases? Evidence from an Analysis of the Relationship between Age and Activities of Daily Living, Comorbidities, and Geriatric Syndromes. Ann Geriatr Med Res. 2024;28(1):65–75.

7. Goeyvaerts N, Hens N, Ogunjimi B, Aerts M, Shkedy Z, Van Damme P, Beutels P. Estimating Infectious Disease Parameters from Data on Social Contacts and Serological Status. Journal of the Royal Statistical Society Series C: Applied Statistics. 2009;59(2):255–77.

8. Palumbo N, Khattab S, Lawson A, Rochon PA. The importance of sex and age disaggregated data. J Am Geriatr Soc. 2023;71(7):2339–42.

9. Miljkovic T, Wang X. Identifying subgroups of age and cohort effects in obesity prevalence. Biometrical Journal. 2021;63(1):168–86.

10. Orosco RK, Hussain T, Brumund KT, Oh DK, Chang DC, Bouvet M. Analysis of age and disease status as predictors of thyroid cancer-specific mortality using the Surveillance, Epidemiology, and End Results database. Thyroid. 2015;25(1):125–32.

